# Obesity- and age-dependent genetic regulation of the plasma proteome in children and adolescents

**DOI:** 10.1101/2025.03.18.25324169

**Authors:** Roman Thielemann, Sara Elizabeth Stinson, Yun Huang, Louise Aas Holm, Justus Florian Gräf, Palle Duun Rohde, Peter Loof Møller, Axel Illeris Poggi, Louise Vølund Anderson, Cilius Esmann Fonvig, Maja Thiele, Aleksander Krag, Simon Rasmussen, Jens-Christian Holm, Torben Hansen

## Abstract

The genetic regulation of the plasma proteome has been extensively studied in adult populations, yet protein quantitative trait loci (pQTL) studies in children and adolescents remain largely unexplored. Here, we mapped pQTLs for 178 plasma proteins measured using affinity-based proteomics in 3,853 Danish children and adolescents (44.1% boys; median age of 11.6 years) from the HOLBAEK Study. We aimed to identify context-dependent pQTLs, where genetic variant-protein associations varied across biological contexts such as obesity, puberty, and sex. We further investigated pQTL variation across the lifespan by comparing our findings to pQTL data from UK Biobank. In the HOLBAEK Study, we identified 1,328 independent, genome-wide significant associations for 178 proteins, replicating previously identified pQTLs in adult cohorts. We identified obesity-dependent *cis*-pQTLs for IL-1ra, TRANCE, and PIgR and found nominally significant evidence for puberty- and sex-dependent genetic differences. Comparative analysis revealed age-dependent differences in *cis*-pQTLs for 30 proteins between children and adolescents of the HOLBAEK Study (aged 4 to 20 years) and adults from the UK Biobank (aged 40 to 70 years). Through one-sample Mendelian randomization analysis, we identified 70 protein–cardiometabolic trait links at nominal significance, highlighting the potential causal role of proteins in cardiometabolic health in children and adolescents.

## Introduction

Plasma proteins represent a molecular intermediate between the genome and the phenome. Protein quantitative trait loci (pQTL) studies have provided valuable insights into the genetic regulation of the plasma proteome in adult populations. With a focus on European adults, large-scale consortia like the UK Biobank and SCALLOP have characterized the genetic regulation of thousands of plasma proteins in tens of thousands of adults using targeted proteomics approaches^1–5^. Studying pQTLs has provided functional insights into known gene-disease associations^4–6^, and combining pQTLs with Mendelian randomization (MR) analysis has identified causal effects of proteins on diseases and traits^2,7^ and supported drug target discovery^2^. Recent studies have also extended our understanding of blood pQTLs in non-European adults^8–10^ and identified pQTLs using untargeted proteomics approaches^9,11^. Overall, pQTL studies have provided a rich genetic basis for functional and causal studies with actionable and translatable insights into the role of plasma proteins in health and disease.

Plasma protein levels are highly correlated with demographic factors such as age, sex, and body mass index (BMI)^1,12^. Emerging evidence has highlighted pQTL differences with sex and age, but studies on pQTL changes with puberty and obesity are sparse^11^. Sex-dependent pQTL differences were identified for a relatively small fraction of plasma^13^ and brain proteins^14^ and both sex and age were found to influence gene-environment interactions of pQTLs^15^. Age-dependent changes were demonstrated for BMI-associated genetic variants, using data collected at several time points covering infancy, childhood, adolescence, and adulthood^16^. In the same study, the performance of polygenic risk scores for BMI varied between age groups, further highlighting the importance of context-dependent genetic effects across the lifespan. For pQTLs, age-dependent differences have been reported in adults from the UK Biobank^1^, but pQTL data from children and adolescents is lacking.

We previously reported mass-spectrometry (MS)-based pQTL data for 1,216 proteins in 2,147 children and adolescents from the HOLBAEK Study, which consisted of two cohorts of Danish children and adolescents: 1) an obesity clinic and 2) a population-based cohort^11^. MS can capture a diverse spectrum of proteins in blood plasma, but low-abundant proteins, such as inflammation markers, remain difficult to measure using MS-based methods. Targeted proteomic approaches, like the antibody-based proximity extension assay, can capture even low-abundant plasma proteins^17^. Pediatric obesity is linked to systemic inflammation, which drives obesity-related cardiometabolic disease^18,19^. Since the cardiovascular disease process begins early in life, early detection and preventive measures in children and adolescents are essential^20,21^. Measuring inflammation and cardiovascular-related plasma proteins provides an opportunity to identify markers of cardiometabolic risk in pediatric cohorts and to study their genetic regulation in early life^12^. Genetic variants associated with protein levels can be leveraged in Mendelian randomization analysis, to identify proteins with causal links to cardiometabolic health^2,3,22^.

Here, we mapped pQTLs for 178 inflammation- and cardiovascular-related plasma proteins measured with Olink proteomics in 3,853 Danish children and adolescents with and without obesity. We explored the interplay between pQTLs and obesity, puberty, age, and sex in children and adolescents and leveraged data from the UK Biobank to compare pQTL effects across age groups. Lastly, we performed Mendelian Randomization using individual level data to identify proteins with potential causal links to cardiometabolic traits in childhood and adolescence.

## Results

### Discovery of pQTLs in children and adolescents

We performed genetic studies of plasma proteins in 3,853 Danish children and adolescents from the HOLBAEK Study (Figure 1). The study cohort consisted of 1) a population-based cohort (n=2,095), recruited from Danish schools^23^ and 2) an obesity clinic cohort (n=1,758), which followed a childhood obesity management program at Holbæk Hospital^24^. Study characteristics of both cohorts were reported in Table S1. We mapped pQTLs for up to 7.7 million imputed genetic variants with minor allele frequencies (MAF) above 0.01 to 178 cardiovascular- and inflammation-related proteins, measured in plasma with affinity-based Olink Target 96 panels at baseline (Cardiovascular II, Inflammation; Table S2). We performed genome-wide association studies (GWAS) on plasma protein levels in the population-based cohort and the obesity clinic cohort, followed by a meta-analysis to map pQTLs in 3,853 children and adolescents.

**Figure 1:**
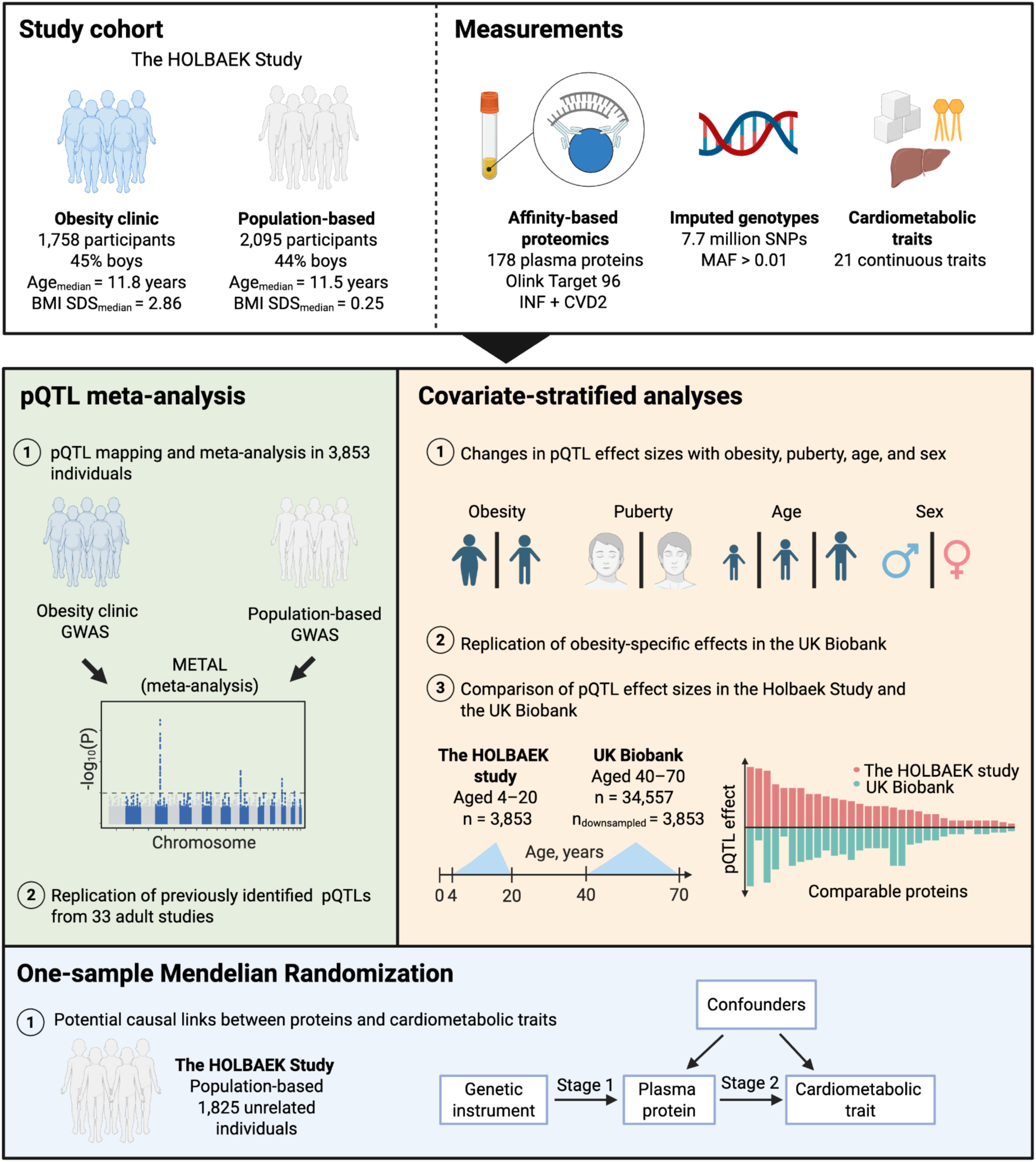
Study overview. Top panel: the two cohorts of the HOLBAEK study and relevant measurements. Middle left panel: pQTL identification in 3,853 children and adolescents. Middle right panel: covariate-stratified analysis in children and adolescents and comparison with the UK Biobank. Bottom panel: one-sample MR analysis between proteins and cardiometabolic traits in the HOLBAEK Study.

We identified 943 primary associations at a study-wide significance level (P < 2.81 × 10^-10^; Table S3) for 118 proteins and 1,328 primary associations at genome-wide significance (P < 5 × 10^-8^) for 138 proteins. We defined primary pQTLs as the most significant variant within a 1 Mb region after clumping (r^2^ > 0.2). Unless specified otherwise, we performed our analyses on study-wide significant pQTLs. *Cis*-pQTLs constituted 713 of 943 (76%) primary associations and were found for 90 of 178 proteins. *Cis*- and *trans-*pQTLs were distributed unevenly across chromosomes, with the highest number of pQTLs on chromosome eleven and the lowest number of pQTLs on chromosomes seven and twenty-two (Figure 2A,B). We observed the strongest effect sizes for pQTLs with low minor allele frequencies (MAF; Figure 2C). We identified at least one study-wide significant *trans*-pQTLs for 63 proteins and ten or more *trans*-pQTLs for 6 proteins (Figure 2D). Likewise, 26 proteins had 10 or more *cis*-pQTLs. Most pQTLs were located in non-coding regions of the genome, with 4% of pQTLs being missense variants, 1% synonymous variants, and <1% stop-gain variants (Figure 2E). We further tested if pQTLs identified in children and adolescents were previously reported in 33 adult pQTL studies (Table S4). All study-wide pQTLs identified in our study were located within 1Mb of previously reported pQTLs for the same protein in adults, underlining the overall similarity of pQTL loci in children and adults.

**Figure 2:**
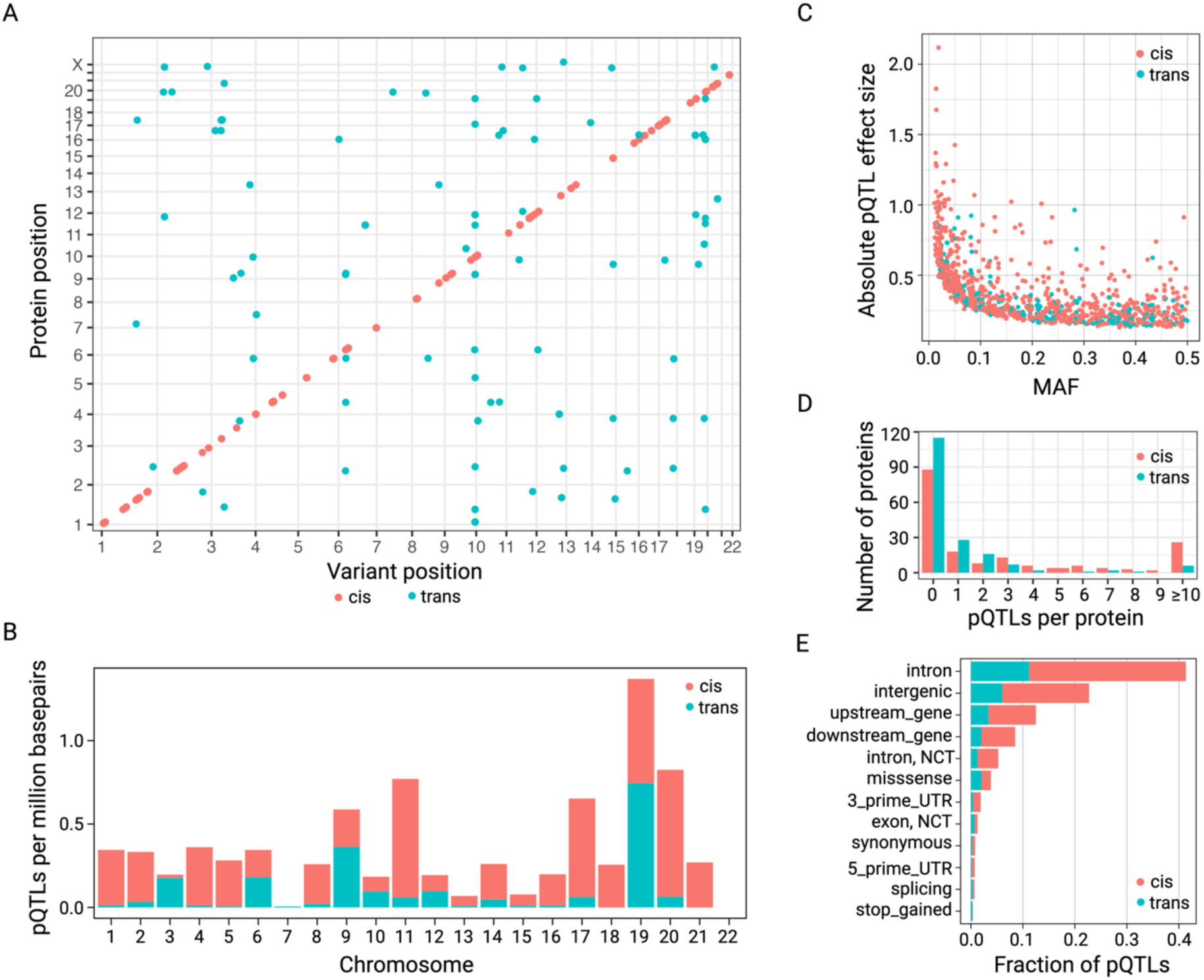
Genetic regulation of proteins in children and adolescents from the HOLBAEK Study. A) Distribution of study-wide significant pQTLs and the transcription start sites of associated proteins across the genome. Results are focussed on autosomal variants. B) Distribution of *cis*- and *trans*-pQTLs across autosomal chromosomes. The number of pQTLs was adjusted for chromosome length and reported as pQTLs per million base pairs. C) Absolute values of GWAS effect sizes plotted against MAF for *cis-* and *trans*-pQTLs. The highest effect sizes were observed in *cis*-pQTLs with low allele frequencies. D) Number of study-wide, independent *cis*- and *trans*-pQTLs per protein. E) Annotations of study-wide *cis*-pQTLs with the Ensembl Variant Effect Predictor (VEP; NCT = non-coding transcript, UTR = untranslated region).

### The genetic regulation of plasma proteins is altered in pediatric obesity

We identified obesity-dependent pQTLs by comparing lead *cis*-pQTLs between the population-based (n=2,095) and the obesity clinic cohorts (n=1,758). Overall, pQTL statistics were similar between the two cohorts: We identified 373 study-wide significant pQTLs for 93 proteins in the population-based cohort after clumping and 316 for 91 proteins in the obesity clinic cohort (Figure 3A). Single nucleotide polymorphism (SNP) effects (*i.e.*, beta coefficients) of the 76 study-wide *cis*-pQTLs agreed in their effect direction without exception (Figure 3B, Table S5). *Cis*-pQTLs explained 0.2% to 40.7% of the variance in protein levels after adjustment for covariates, with a median of 6.16% and 5.75% of variance explained in the population-based and obesity clinic cohorts, respectively (Figure 3C). We identified significant obesity-dependent differences in *cis*-pQTLs effect sizes (FDR < 5%) for four proteins: PIgR, IL-1ra, TRANCE, and CXCL10 (Figure 3B). *Cis*-pQTLs of PIgR, IL-1ra, and TRANCE explained a low proportion of variance in protein levels in the population-based cohort (below 3.2%), but up to 8.3% in individuals from the obesity clinic (Figure 3C). We further characterized the obesity-dependent loci by performing gene-environment interaction analysis with BMI standard deviation score (SDS). All four loci showed nominally significant interaction with BMI SDS, with the most significant interaction for IL-1ra (P-value=8.01 × 10^-6^) and less significant interaction for TRANCE (P-value=6.60 × 10^-3^). We excluded CXCL10 from further analyses because of a sparse LD structure in the region surrounding the lead SNP, indicating a potentially spurious GWAS signal (Figure S1). Together, these data demonstrate an interaction between pediatric obesity and the genetic regulation of PIgR, IL-1ra, and TRANCE.

**Figure 3:**
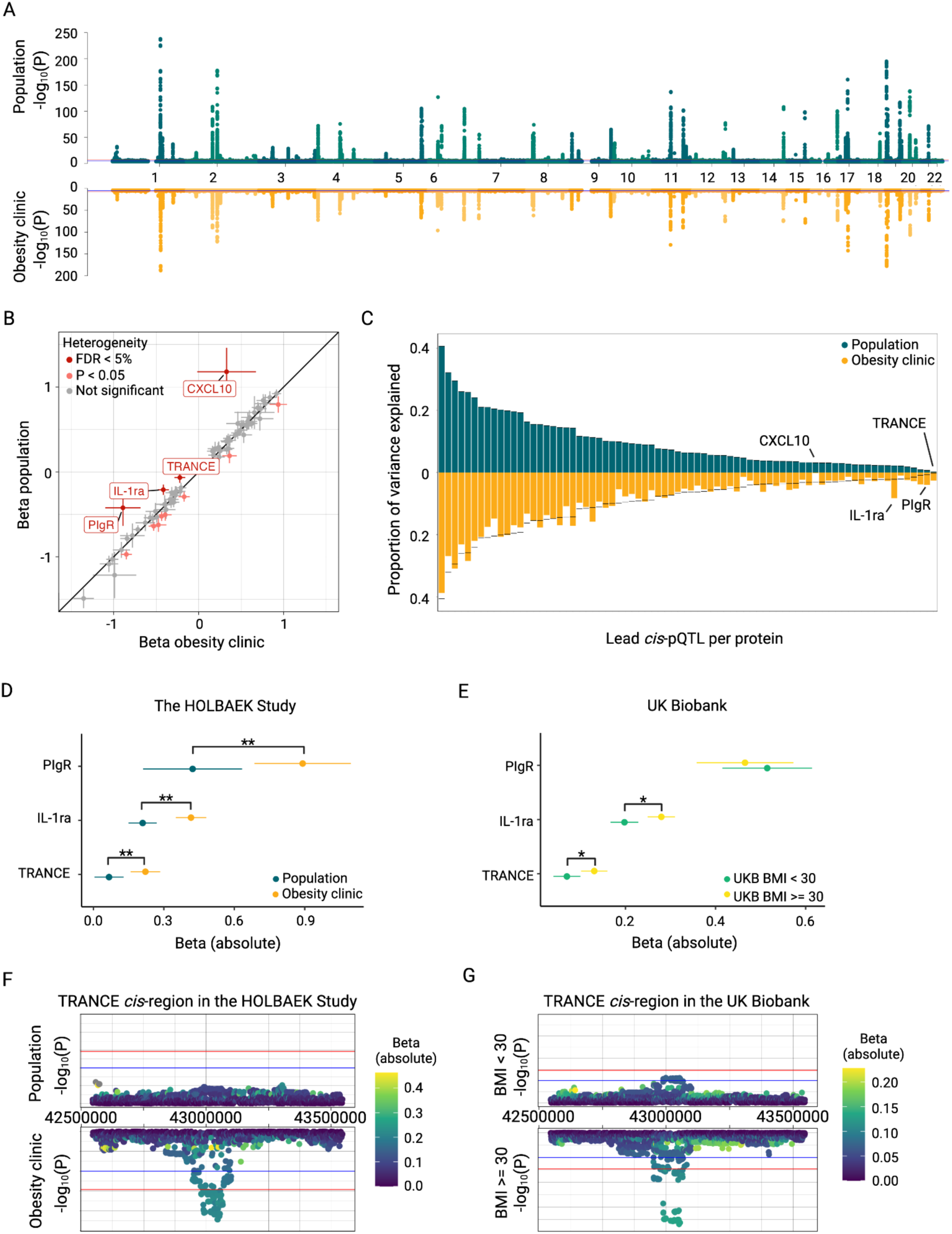
*Cis*-pQTLs differences in population-based and obesity clinic cohorts. A) GWAS signals for 178 distinct plasma proteins in children and adolescents from the population (n = 2,095) and obesity clinic (n = 1,758). B) Differences in beta values of plasma *cis*-pQTLs across the population and obesity clinic cohorts. We found evidence for obesity-dependent heterogeneity (FDR < 5%) in c*is*-pQTLs for PIgR, IL-1ra, TRANCE, and CXCL10. C) *Cis*-pQTLs explained a similar proportion of variance in protein levels across population and obesity clinic cohorts. Proteins with evidence for obesity-specific heterogeneity were labeled. D) *Cis*-pQTL effect sizes for PIgR, IL-1ra, and TRANCE differed in the obesity clinic and population-based cohorts of the HOLBAEK Study. The double asterisk marks evidence for obesity-dependent heterogeneity (FDR < 5%) in the meta-analysis. E) Obesity-dependent differences replicated for IL-1ra and TRANCE in adults with a BMI ≥ 30 kg/m^2^ (n=8,803) and a control group with BMI < 30 kg/m^2^ (n=8,803) from the UK Biobank. The asterisk marks nominal significant heterogeneity in the meta-analysis. Zoom plots of the *cis*-region for TRANCE revealed obesity-dependent differences in both F) the HOLBAEK Study and G) the UK Biobank. Suggestive significance (10^-5^, blue) and genome-wide significance (5 × 10^-8^, red) thresholds are highlighted. Genome

### Obesity-dependent pQTL differences persist into adulthood

Next, we aimed to characterize the source of obesity-dependent heterogeneity in *cis*-pQTLs. We found that inverse rank transformed (INT) protein levels were increased in the obesity clinic cohort for IL-1ra, TRANCE, and decreased for PIgR compared to the population-based cohort (Figure S2A, Table S6). We found no genome-wide significant association of *cis-*loci with childhood BMI SDS in GWAS summary statistics from 39,620 individuals of the EGG consortium^25^. We conducted PheWAS using the ‘otargen’ R package^26^ and found significant phenotype associations for lead *cis*-pQTLs of IL-1ra and TRANCE (Table S7). For TRANCE, the protein-increasing allele was strongly associated with bone density measures, including increased heel bone mineral density and decreased prevalence of osteoporosis, as well as with decreased prevalence of hypothyroidism. The IL-1ra *cis*-pQTL was inversely associated with decreases in immunological measures, such as c-reactive protein levels and neutrophil counts.

To replicate obesity-dependent pQTLs in adults, we performed an obesity-stratified pQTL analysis in the UK Biobank. We identified pQTLs for PIgR, IL-1ra, and TRANCE in 8,083 UK Biobank participants with a BMI ≥ 30 kg/m^2^ and compared them to pQTLs identified in a sex-matched control group consisting of 8,083 UK Biobank participants with a BMI < 30 kg/m^2^. Out of the three proteins with obesity-dependent pQTLs in the HOLBAEK Study (Figure 3D), we replicated obesity-dependent differences (heterogeneity P-value < 0.05) for the lead *cis*-pQTLs of IL-1ra and TRANCE in the UK Biobank (Figure 3E). Obesity-dependent signals for PIgR did not replicate. The UK Biobank and HOLBAEK Study proteomics data have been generated using the Olink Explore 3072 panel and the Olink Target 96 panel, respectively. In an independent cohort of 739 individuals, we found high correlation across platforms for levels of TRANCE and IL-1ra (Pearson ρ > 0.8; Table S8) but lower correlation for PIgR (Pearson ρ = 0.32). Notably, *cis*-pQTLs for TRANCE reached genome-wide significance in individuals with obesity in both the HOLBAEK Study (Figure 3F) and the UK Biobank (Figure 3G), but not in the control groups with identical or higher sample sizes. Protein levels were increased in individuals with obesity for PIgR, IL-1ra, and TRANCE in the UK Biobank (Figure S2B, Table S6).

### Differences in *cis*-pQTLs before and after puberty

We then investigated the influence of puberty on the genetic regulation of proteins. First, we performed puberty-stratified GWAS in a subset of individuals with recorded Tanner stage, a measure of pubertal progression, and compared pQTLs between prepubertal (n=1,091) and pubertal/post-pubertal (n=1,731) individuals. Overall, pQTL statistics in individuals before and during/after puberty were highly comparable. We identified study-wide significant *cis*-pQTLs for 68 proteins in the pubertal/post-pubertal group. Effect sizes of *cis*-pQTL generally agreed between the two groups (Figure 4A, Table S9). We found nominally significant puberty-dependent differences in *cis*-pQTLs for five proteins (Figure 4A), of which differences for LOX1 and TM were the most significant (heterogeneity P-value 2.70 × 10^-3^ and 5.60 × 10^-3^). LOX1 and TM *cis*-pQTLs reached study-wide significance in both groups, but effect sizes were significantly higher in the prepubertal group compared to the pubertal/post-pubertal group. Lead *cis*-pQTLs for LOX1 and TM explained 6.9% and 10.8% of variance in the prepubertal group but only 2.5% and 5.4% of variance in the pubertal/post-pubertal group. PheWAS analysis for the two lead *cis*-pQTLs did not reveal significant associations, besides the *cis*-pQTL for TM being associated with TM levels^26^.

**Figure 4:**
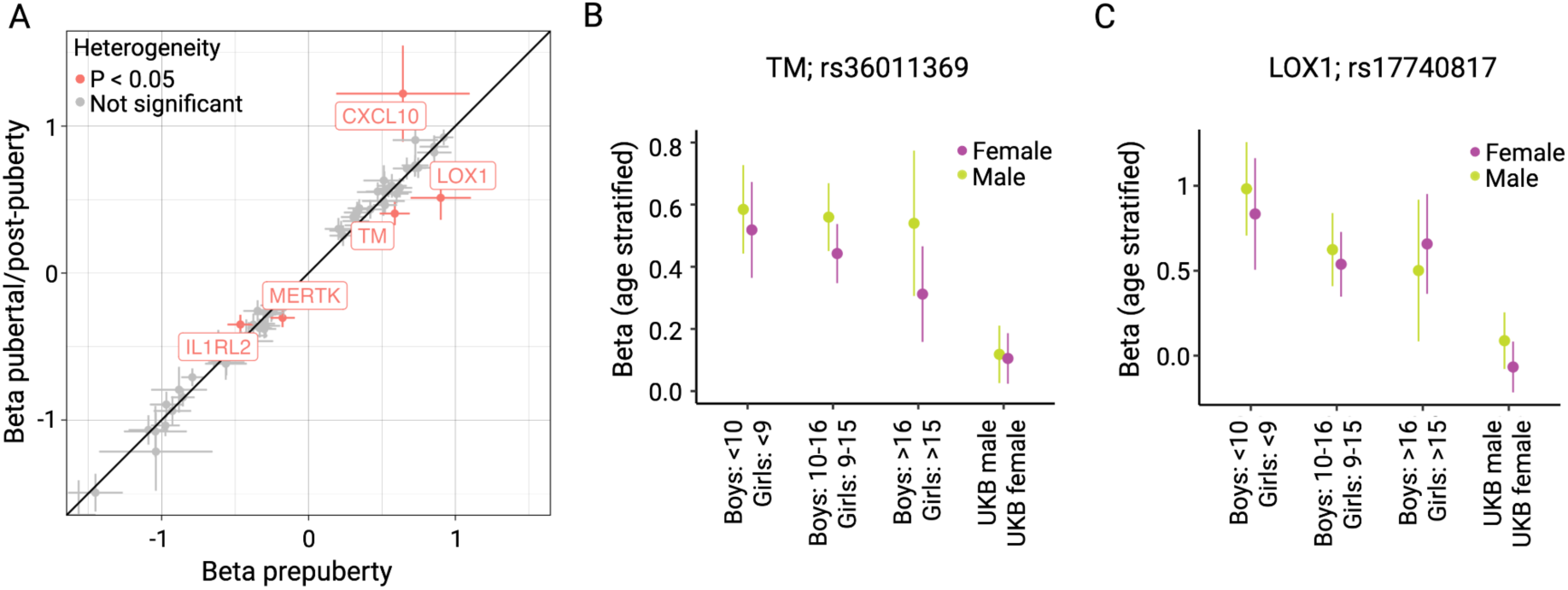
Puberty- and age-dependent *cis*-pQTL differences in the HOLBAEK Study. A) Nominally significant differences in beta values of plasma *cis*-pQTLs between prepubertal (n=1,091) and pubertal/post-pubertal (n=1,731) individuals in the HOLBAEK Study. Puberty-dependent differences were most significant for LOX1 and TM. Age (in years) trajectories of effects sizes for B) TM and C) LOX1 *cis*-pQTLs for boys and girls in the HOLBAEK Study and a subset of the UK Biobank European discovery cohort. The HOLBAEK study was stratified into the three age groups by sex: girls below 9 years (n=487), girls between 9 to 15 years (1,218), and girls above 15 years (n=447), as well as boys below 10 years (n=579), boys between 10 to 16 years (n=927), and boys above 16 years (n=195). Sex-stratified GWAS in the UK Biobank were performed in a subset of 1,218 females and 927 males.

We further assessed how associations of LOX1 and TM *cis*-pQTLs changed with age, independent of Tanner stage. We separated the HOLBAEK Study into three age groups based on normative Danish references for pre-puberty, puberty, and post-puberty and ran sex-stratified pQTL analysis in adults of the UK Biobank to compare *cis*-pQTLs for LOX1 and TM across the lifespan. Notably, sample sizes varied across age groups. Cross-platform correlations of protein measurements between the Olink Target 96 panel and the Olink Explore 3072 panel were moderate for LOX1 and TM (Pearson ρ=0.78 and 0.57; Table S8). We found decreased beta values for LOX1 in boys aged 10–16 compared to those under 10 (heterogeneity P-value < 0.05), while other differences among age groups in the HOLBAEK Study were not significant (Figure 4B, C; Table S10). Additionally, beta values for TM and LOX1 were decreased in adults of the UK Biobank compared to youths in all three age groups of the HOLBAEK Study, except for LOX1 in boys over 16 (heterogeneity P-value < 0.05).

### pQTLs differed across the lifespan

To systematically assess pQTL differences between youth and adults, we compared lead *cis*-pQTLs between the HOLBAEK Study (n=3,853; 4 to 20 years) and a subset of the UK Biobank European ancestry discovery cohort (n=34,557; 40 to 70 years). First, we investigated whether differences in sample size, covariates used in the association analysis, and GWAS methods (GEMMA vs. REGENIE) could bias the comparison across cohorts. Rerunning the association analysis in the HOLBAEK Study with REGENIE and covariates comparable to the UK Biobank led to a median decrease in beta estimates of 0.039 (Q1–Q3: 0.028–0.062) across *cis*-pQTLs (Figure S3). Similarly, downsampling the UK Biobank cohort from 34,557 to 3,853 individuals to ensure comparable sample sizes between cohorts systematically decreased beta estimates by 0.051 (Q1–Q3: 0.022–0.087) in the UK Biobank (Figure S3). These systematic shifts in beta values affected the number of proteins with age-dependent *cis*-pQTL differences. We initially identified 38 proteins with age-dependent differences (FDR < 5%), which decreased to 30 proteins after matching covariates and sample sizes. Despite this difference, 27 proteins with significant differences were shared between the two analyses.

We identified age-dependent differences in *cis*-pQTLs for 30 proteins between the HOLBAEK Study and the UK Biobank, including LOX1 and TM (Figure 5A and Figure S4; Table S11). *Cis*-pQTLs with age-dependent differences in beta values explained between 1% and 17.6% of the variance in protein levels in the HOLBAEK Study (Figure 5B). The largest differences in explained variance were found for CD8A (3.1% in UK Biobank and 17.6% in HOLBAEK; Figure 5C) and FGF5 (15.2% in UK Biobank and 3.9% in HOLBAEK; Figure 5D). Besides these differences, overall pQTL statistics for the UK Biobank and the HOLBAEK Study were similar: allele frequencies of *cis*-pQTLs were highly correlated (Pearson ρ=0.999 and *cis*-pQTL effect sizes correlated well between the two cohorts (Pearson ρ=0.952). After addressing differences in sample size, covariates, and GWAS methods, we identified age-dependent pQTL differences in 30 of 75 proteins with shared study-wide *cis*-pQTLs, suggesting changes of the genetic regulation of the plasma proteome across the lifespan.

**Figure 5:**
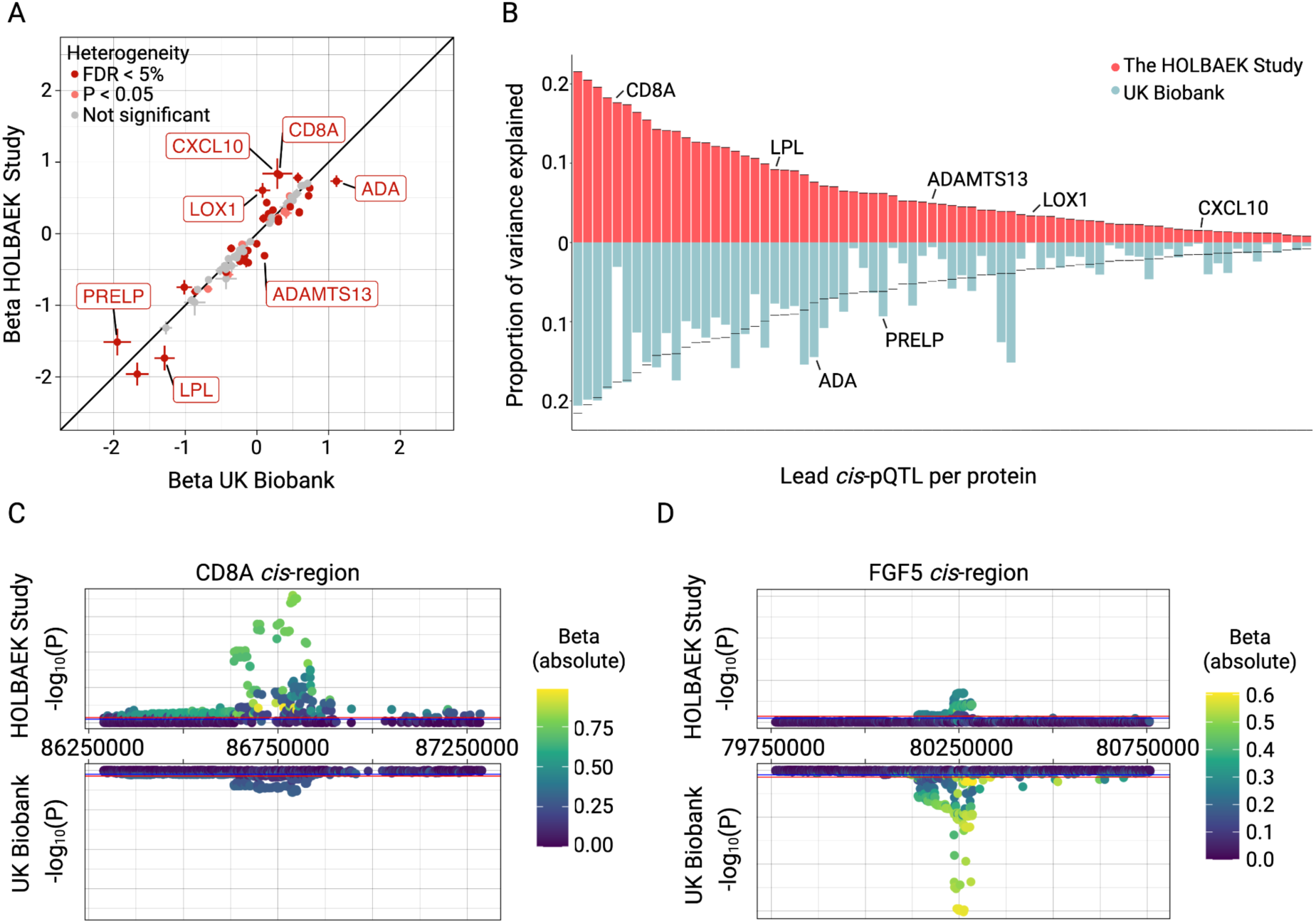
*Cis*-pQTL differences between children and adolescents in the HOLBAEK Study and adults in the UK Biobank. A) Differences in effect sizes of plasma *cis*-pQTLs across the HOLBAEK Study (n=3,853; 4 to 20 years) and the downsampled UK Biobank (n=3,853; 40 to 70 years). We found significant differences in c*is*-pQTLs (FDR < 5%) for 30 of 75 compared proteins. Labels were added for proteins with an absolute difference in beta values over 0.3. B) Proportion of variance explained by *cis*-pQTLs across cohorts. Annotated proteins from Figure 5A were highlighted. Zoom plots of *cis*-regions for C) CD8A and D) FGF5 revealed age-dependent pQTL differences between the HOLBAEK Study and the UK Biobank. Genomic coordinates in this figure refer to genome-build GRCh38. Suggestive significance (10^-5^, blue) and genome-wide significance (5 × 10^-8^, red) thresholds are highlighted.

### Age-dependent differences are not driven by differences in proteomic approaches

Next, we asked whether differences in Olink technologies drove pQTL differences between the UK Biobank and the HOLBAEK Study. While the UK Biobank used the Explore 3072 panel with an NGS readout, the Target 96 panels with a qPCR readout were used in the HOLBAEK Study. We compared the correlation of 178 protein levels across the two platforms in 739 individuals without coronary artery disease (CAD) from the Dan-NICAD cohort^27,28^ and found moderate cross-platform correlation (median Pearson ρ=0.69, Q1–Q3: 0.57–0.80; Table S8 and Figure S5). For proteins with study-wide *cis*-pQTLs, we found no difference in correlation between proteins with and without evidence for age-dependent heterogeneity (Pearson ρ=0.713 and 0.736; Wilcoxon P-value=0.69), indicating that age-dependent variation in pQTLs was not primarily driven by differences in Olink technologies. Of the 30 proteins with age-dependent pQTLs, ADAM-TS13 and IL-15RA were the only two with Pearson correlation estimates below 0.5, 11 proteins fell between 0.5 and 0.7, and 17 had estimates above 0.7. We found stronger correlations across Olink platforms for proteins with study-wide *cis*-pQTLs in the HOLBAEK Study (median Pearson ρ=0.73) compared to proteins without identified *cis*-pQTLs (median Pearson ρ=0.64; Wilcoxon P-value=7.74 × 10^-5^).

### Nominal Evidence of sex-dependent pQTL differences in children and adolescents

Next, we assessed sex-dependent pQTL differences in 1,701 boys and 2,152 girls of the HOLBAEK Study. Overall, pQTL loci and effect sizes agreed between boys and girls (Figure S6, Table S12). We found two study-wide significant loci in boys that did not reach genome-wide significance in girls: a *cis*-pQTL for FGF21 (rs838131) and a *trans*-pQTL for VSIG2 (rs708686). Although not genome-wide significant, these loci reached suggestive significance (P-value < 10^-5^) in girls, indicating that they might be captured in both sexes with higher sample sizes. We compared the effect sizes of lead *cis*-pQTLs between boys and girls for 77 proteins and found only nominally significant evidence for TM, CXCL1, and IL1RL2. In UK Biobank adults, Koprulu et al. found sex-dependent pQTL differences in 3.9% of proteins (Olink Explore 1536), but pQTLs for TM, CXCL1, and IL1RL2 were not significant^13^.

### Causal relationships between proteins and cardiometabolic traits in children and adolescents

We performed one-sample MR using two-stage least squares analysis to identify potentially causal protein-cardiometabolic trait links in 1,825 children and adolescents from the population-based cohort. We included 90 proteins with study-wide significant *cis*-pQTLs and 21 cardiometabolic traits in the analysis. We identified 70 protein-cardiometabolic trait pairs with nominally significant associations (Figure 6A,B; Figure S7). Only one protein, MMP7 passed multiple testing correction. All nominally significant associations had F-statistics > 10, indicating no weak-instrument bias (Table S13). Few proteins were exclusively associated with cardiometabolic traits from a single category, including MMP7, MERTK and MMP with liver-related traits and ENRAGE with blood pressure traits (Figure 6B,C). Several proteins were associated with cardiometabolic traits from different categories, such as IL15RA (Figure 6D), indicating a complex link between protein levels and cardiometabolic traits in children and adolescents.

**Figure 6:**
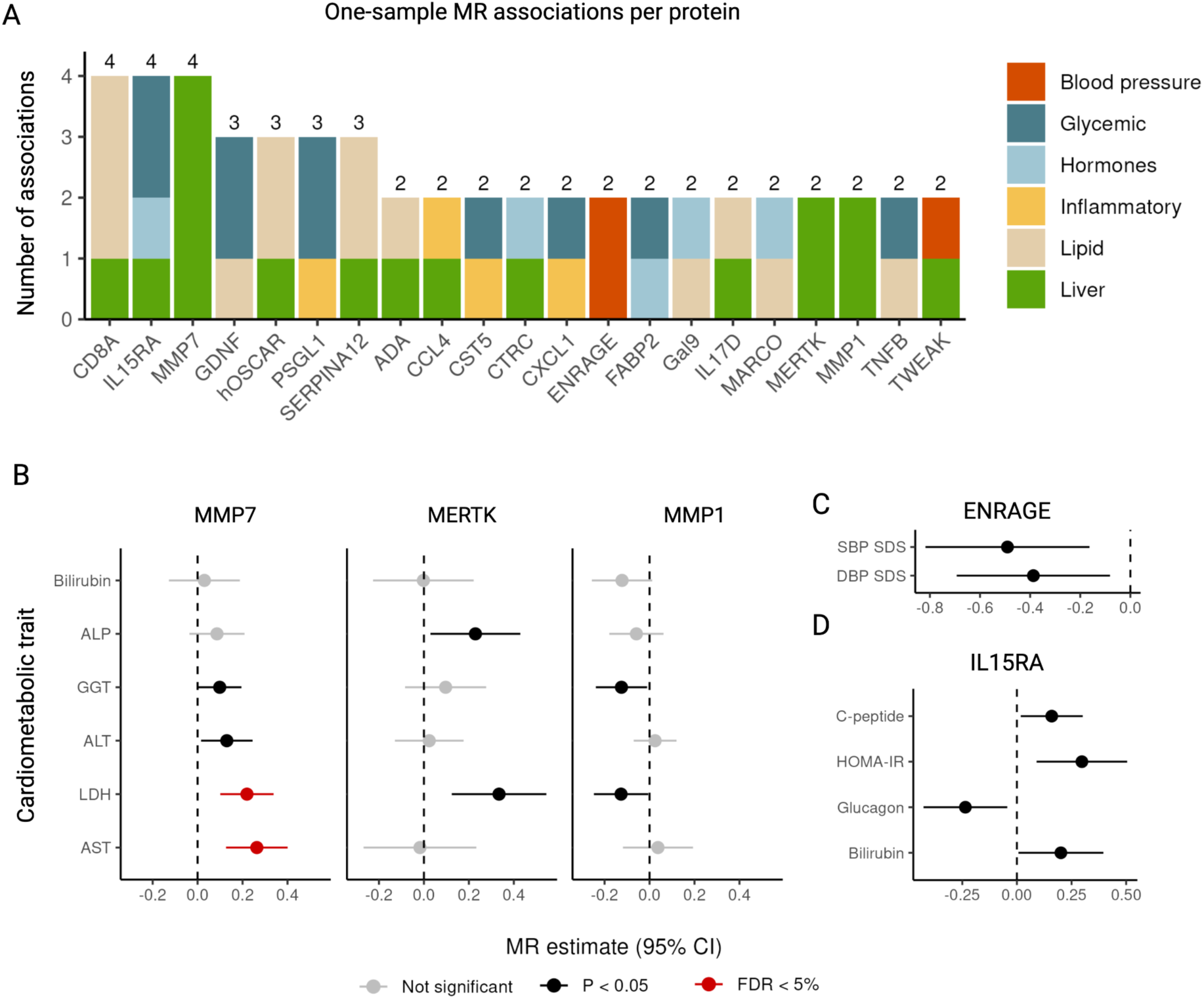
One-sample MR analysis links plasma proteins to cardiometabolic traits in children and adolescents. A) Overview of plasma proteins with at least two associations with cardiometabolic traits. Cardiometabolic traits were grouped into six categories. B) Associations of MMP7, MERTK, and MMP1 with liver-related traits. C) Associations of ENRAGE with blood pressure. D) Associations of IL15RA with glycemic traits.

## Discussion

The genetic regulation of plasma proteins has been thoroughly studied in adult populations, but pQTL studies in children and adolescents are sparse. Here, we presented findings from a pQTL study of 178 cardiovascular- and inflammation-related proteins in 3,853 Danish children and adolescents (aged 4 to 20 years). We reported 1,328 primary associations at genome-wide significance level for 178 targeted plasma proteins and replicated previously identified pQTLs from adult cohorts. We explored the interplay of *cis*-pQTLs with obesity, puberty, and sex in children and adolescents. We identified obesity-dependent differences in *cis*-pQTLs of PIgR, IL-1ra, and TRANCE (FDR < 5%) and found nominally significant evidence for puberty- and sex-dependent genetic differences. Lastly, we leveraged pQTL data from the UK Biobank to identify differences in pQTLs across the lifespan, and to replicate context-dependent differences in an adult cohort. We found age-dependent differences between children and adolescents from the HOLBAEK Study and adults from the UK Biobank in *cis*-pQTLs of 30 proteins. We further replicated obesity-dependent findings for IL-1ra and TRANCE in individuals from the UK Biobank. Lastly, we identified several proteins with potential causal links to cardiometabolic traits in childhood and adolescence.

The number of large-scale pQTL studies in children and adolescents is limited: in the HOLBAEK Study, pQTLs were previously reported for 1,216 plasma proteins measured with mass spectrometry in 2,147 individuals^11^. In the ALSPAC cohort, 92 plasma proteins were measured with the Olink Target Inflammation panel in 3,000 9-year-olds, but pQTL data was not yet published^29^. Here, we contributed with pQTL data for 178 proteins measured in 3,853 Danish 4–20-year-olds. The HOLBAEK Study included both MS-based^11^ and targeted plasma proteomic data^12^ with complementary information. Seven proteins were captured by both methods. The MS-based approach captures a broader spectrum of relatively abundant proteins in blood plasma and provides additional information on peptide abundance. The affinity-based Olink approach captures a smaller set of disease-related and potentially low-abundant proteins in the HOLBAEK Study. Affinity-based plasma proteomic data is currently available in several cohorts and biobanks such as the UK Biobank, SCALLOP, the HOLBAEK Study, and ALSPAC and provides exciting opportunities to compare pQTLs across cohorts and age ranges^1,2,29^.

We leveraged the unique characteristics of the HOLBAEK Study, such as the focus on children and adolescents and the recruitment from population- and obesity clinic-based cohorts, to perform puberty-, age-, and obesity-stratified analyses not feasible in other cohorts. These analyses were in part motivated by a study by Helgland et al. that identified age-dependent changes in effect size for BMI-associated variants and highlighted their importance for the performance of polygenic risk scores^16^. Similar to Helgeland’s findings for the interplay between genetic variants and BMI, our findings indicate that genetic associations with protein levels differ across the lifespan. Not only did we find pQTLs to differ significantly with age, but also with obesity, indicating that the genetic regulation of protein levels can be dynamic and dependent on biological contexts.

We found obesity-dependent *cis*-pQTL differences for IL-1ra and TRANCE in the HOLBAEK Study (Figure 3B) and in the UK Biobank (Figure 3E, F). IL-1ra is a member of the IL-1 family that binds to IL-1 receptors without inducing an intracellular response^30^. IL-1ra has two roles in regulating metabolism: it has anti-inflammatory properties which can prevent hepatic steatosis and atherosclerosis, and it is associated with leptin and insulin resistance^31^. In agreement with the functions of IL-1ra, we found the protein-increasing allele of its *cis*-pQTL to be inversely associated with immunological measures, such as C-reactive protein and neutrophil counts. Evidence from MR studies suggested a protective role of IL-1ra levels for rheumatoid arthritis and increasing effect on total cholesterol^2^. TRANCE, or TNFSF11, is involved in osteoclast differentiation and activation, and regulates the T-cell-dependent immune response^32–34^. TRANCE is known for driving bone loss in osteoporosis and a therapeutic target for bone-related diseases^32,35–38^. In contrast, genetic findings point towards a protective effect of TRANCE plasma levels on bone loss. The protein-increasing allele of the TRANCE *cis*-pQTL positively associated with heel bone mineral density, and negatively associated with the prevalence of osteoporosis and hyperthyroidism. Evidence from MR studies indicated that TRANCE is positively associated with bone mineral density and age at menarche and negatively associated with bone fractures and primary biliary cirrhosis^2^. Taken together, plasma levels of proteins affecting inflammation-related traits may be subject to increased effects of genetic regulation in both children, adolescents, and adults with obesity.

Several Mendelian Randomization studies have linked proteins to cardiometabolic outcomes in adults, using both two-sample MR^2,3^ and one-sample MR approaches^22^. Here, we performed one-sample MR using individual-level data and identified potential causal protein-cardiometabolic trait links in children and adolescents. Matrilysin (MMP7), a metalloprotease has previously been linked to tissue remodelling in liver and pulmonary fibrosis in adults^39–41^. Our findings suggest a potentially causal link between MMP7 and markers of liver health, already in childhood and adolescence. Other nominally significant protein-cardiometabolic trait associations observed in this study also have known links to cardiometabolic health in adults. This includes MMP1 and MERTK, which were linked to liver-related traits in our study and are known as potential drug targets for liver fibrosis^42,43^. ENRAGE is a known marker for atherosclerosis and incident coronary heart disease^44,45^ and was linked to blood pressure in our study.

Despite being one of largest pQTL studies in children and adolescents to date, the sample size of this study is much smaller than previous large-scale pQTL studies in adults (e.g., SCALLOP, UK Biobank). While pQTLs, especially *cis*-pQTLs, can be robustly identified at smaller sample sizes^11,46^, stratifying individuals based on covariates further reduces the sample size, decreasing the power to detect context-dependent pQTL differences. Gene-environment interaction analysis is often used to capture the interaction of non-genetic environmental factors (like BMI SDS) with genetics, but performing such analyses on a genome-wide scale requires large sample sizes, and preselection of variants of interest is often necessary^47^. Given the sample size constraint of this study, we prioritized study-wide significant *cis*-pQTLs in our covariate-stratified comparison and the interaction analysis with BMI SDS. Another limitation is the lack of pediatric cohorts for replication. We used data from the UK Biobank to replicate identified pQTLs and obesity-dependent pQTL differences. Using MS-based pQTL data for replication is hindered by the small overlap of proteins (n=7) between our study and the MS-based pQTL study^11^. Future pQTL studies in pediatric cohorts, like the ALSPAC cohort^29^, and meta-analysis of their results will provide broader and more robust insights and greatly benefit the field.

Comparing pQTLs between two cohorts such as the HOLBAEK Study and the UK Biobank is challenging. We aimed to identify age-dependent pQTL differences, but cohort differences other than age—such as medications, comorbidities, and selection criteria—can bias this comparison and result in pQTL differences that are falsely attributed to age. We addressed technical differences between the cohorts that could bias association analysis results such as differences in sample size, covariates, GWAS methods, and proteomics methodology. We found that proteomic cross-platform correlation was not significantly decreased for proteins with age-dependent pQTLs, indicating that Olink platform differences were not the primary driver of age-dependent pQTL differences. Cross-platform correlation estimates for 178 proteins in the Dan-NICAD cohort were moderate (median Pearson ρ=0.69) when compared to previously reported correlation estimates for proximity extension assays using NGS or qPCR readouts^48^ (median Pearson ρ=0.985). Some intra- and inter-platform variation in measurements is expected, as a coefficient of variation of 16.5% was reported for the Olink Explore 3072 panel in the UK Biobank^4^. In the Dan-NICAD cohort, Olink Target and Olink Explore data were measured in different laboratories and any differences in sample preparation and handling are expected to contribute to variation across measurements. Differences in proteomics measurements and other sources of bias should be carefully considered when comparing pQTL data across cohorts.

In our study, we observed differences in the effect sizes of SNP-protein associations and gene-environment interactions for SNPs with BMI SDS. However, the cause of these context-dependent differences in pQTLs is not clear. Age- and obesity-dependent differences in SNP-protein associations may result from biological differences in genetic regulation (e.g., chromatin accessibility or gene expression) or from unaccounted interactions of SNPs with environmental factors. Functional follow-up analysis of the identified variants and characterization of molecular changes with age or obesity could help clarify the underlying mechanisms of context-dependent pQTLs.

## Methods

### Study cohort

The study was based on children and adolescents from the HOLBAEK Study, previously known as The Danish Childhood Obesity Biobank^49^. Two groups of children and adolescents were included: 1) an obesity clinic group (n=2,555), which followed a childhood obesity management program at Holbæk Hospital^24^ and included children and adolescents with a BMI ≥ 90th percentile (BMI SDS ≥ 1.28) and 2) a population-based group (n=2,734), recruited from schools across 11 municipalities in Zealand, Denmark^23^. Individuals from both groups were enrolled in the study between August 2007 and April 2019. We excluded individuals 1) with diagnosed diabetes, 2) treated with insulin, statins, liraglutide, or metformin, 3) type 2 diabetes based on blood sample, i.e. fasting plasma glucose > 7.0 mmol/L and/or glycated hemoglobin A_1c_ [HbA_1c_] > 48 mmol/mol), and 4) with a blood sample for proteomic analyses taken more than 90 days apart from clinical measurements. After applying the exclusion criteria, we included 2,095 individuals (44% boys) with a median age of 11.5 years from the population-based cohort and 1,758 individuals (45% boys) with a median age of 11.8 years from the obesity clinic cohort with genotype and plasma proteomic data. Characteristics of both cohorts were reported in Table S1.

### Genomic data processing in the HOLBAEK Study

Genotyping was performed in three batches on the Illumina Infinium HumanCoreExome-12 v1.0 and HumanCoreExome-24 v1.1 Beadchips (Illumina) using Illumina’s HiScan system. Genotypes were called using the Genotyping module (version 1.9.4) of GenomeStudio software (version 2011.1; Illumina). Data were phased using EAGLE2 (version 2.0.5) and imputed using PBWT to the Haplotype Reference Consortium (HRC, r1.1; GRCh37, hg19) on the Sanger server^50^. We removed individuals with more than 5% missing genotypes, too high or low heterozygosity (inbreeding coefficient abs(F) > 0.2), sex mismatches between phenotype and genotype, and European ethnic outliers. We removed SNPs with a call rate below 95% and and those that deviated from Hardy-Weinberg equilibrium at a P-value below 1 × 10^-6^. We included autosomal SNPs with a minor allele frequency above 1% and an imputation score over 0.7. Genomic coordinates refer to genome build GRCh37 unless specified otherwise.

### Proteomic data processing and quality control

Proteomic data was measured with the proximity extension assay technology, using the Olink Target 96 Cardiovascular II panel and the Olink Target 96 Inflammation panels. Protein levels for 178 unique proteins were captured. Six proteins were overlapping between the two panels, in which case only protein measures from the CVD-II panel were included in the analysis. Blood plasma samples were analyzed in three batches. Samples were completely randomized according to subcohort status and observed to have minimal batch effects. Quality control and batch correction were performed using bridge normalization with the OlinkAnalyze R package. Samples that did not pass OlinkAnalyze quality control were removed and values below the limit of detection were kept for GWAS analyses.

### Genetic association analysis in the HOLBAEK Study

GWAS were performed with the additive linear mixed model approach implemented in GEMMA v0.98.3^51^ on imputed genotype dosages. NPX protein levels were rank-inverse normal transformed and linearly adjusted for the year of blood sample collection, Olink panel batch and age, sex, and cohort status, depending on the analysis. The genotyping batch was added as a covariate in the association analysis with GEMMA. Individuals from the obesity clinic and population-based cohort were analyzed separately and results were meta-analyzed using METAL^52^ to obtain summary statistics for all 3,853 participants. METAL was run in the standard error scheme without genomic control correction. Genomic inflation was well controlled with a median genomic inflation factor of 1.003 (Q1–Q3: 0.998–1.006) in the population cohort and 1.003 (Q1–Q3: 0.999–1.007) in the obesity clinic cohort.

The pQTLs passing a multiple-comparison-corrected threshold (P < 2.7 × 10^-10^) after meta-analysis were reported as study-wide significant. Primary associations were defined as study-wide significant variants remaining after clumping (r^2^ > 0.2) in a 1MB region around significant variants using PLINK^53^.

We assessed the novelty of identified study-wide pQTLs by comparing them to pQTLs reported in 33 adult pQTL studies (Table S4) following the approach described by Niu et al.^11^. For all adult studies, we retained the pQTLs at the reported significance levels. We defined a pQTL as novel if no variants within 1Mb of our primary pQTL had been previously associated with the corresponding protein. We did not consider linkage disequilibrium in this analysis.

### Variant annotation

We annotated pQTLs using the Ensembl Variant Effect Predictor (VEP) version 111 with the “--pick” option^58^. *Cis*-pQTLs were defined as pQTLs within 1 million base pairs of the transcription start site. We mapped UniProt IDs to Ensembl BioMart Release 113^59^ (GRCh38) to identify transcription start sites, prioritizing canonical transcripts on primary chromosomes. For hOSCAR—whose canonical transcript was not located on a primary chromosome—we used the start site of the longest protein-coding transcript. Genomic coordinates were lifted over to GRCh37 with the liftover R package^61^.

### PheWAS analysis

We conducted PheWAS analysis using the ‘otargen’ R package^26^. We included SNP-trait associations from FinnGen, UK Biobank, and the GWAS catalog with a P-value below 0.05 divided by 3,618, the total number of GWAS studies where the variant is associated.

### Covariate-stratified analysis

Covariate-stratified analyses were performed for obesity, sex, age, and Tanner stage, a measure of pubertal progression ranging from one to five. Protein levels (NPX values) were rank-inverse normal transformed and covariate-adjusted separately in each stratified cohort. Obesity-stratified GWAS were performed on individuals from the population (n=2,095) and obesity clinic (n=1,758) and protein levels were additionally adjusted for age and sex. Sex-stratified GWAS were performed in boys (n=1,701) and girls (n=2,152) with additional adjustments for age and cohort status. The pubertal status of 2,822 participants was assessed using Tanner stages^54,55^. Tanner stage-stratified GWAS were performed in prepubertal (n=1,091; Tanner stage 1), and pubertal/post-pubertal participants, (n=1,731; Tanner stage 2–5) with additional adjustment for cohort status and sex. Lastly, age-stratified analyses were performed for girls below 9 years (n=487), girls between 9 to 15 years (1,218), and girls above 15 years (n=447), as well as boys below 10 years (n=579), boys between 10 to 16 years (n=927), and boys above 16 years (n=195). Stratification into age groups was based on data on pubertal development in Danish children^56^.

We compared effect sizes and the proportion of variance explained by lead *cis*-pQTL variants for each protein. Lead *cis*-pQTLs were defined as the *cis*-pQTL variant explaining the highest proportion of variance after clumping with PLINK. We included lead *cis*-pQTLs that achieved study-wide significance in one or both of the stratified cohorts for the comparison. The proportion of variance explained was calculated from the GWAS effect sizes (beta), standard error of beta (se), and sample size using the formula: proportion of variance explained = beta^2^ /(beta^2^+se^2^*N). Heterogeneity in obesity-, puberty-, and sex-stratified GWAS was assessed with METAL^52^ and heterogeneity in sex-stratified age groups was assessed with a fixed-effects model implemented in the metafor R package^57^. Error bars spanned the 95% confidence interval unless indicated otherwise.

### Gene-environment interaction analysis

We performed gene-environment interaction analysis with BMI SDS in 3,853 individuals of the HOLBAEK Study using GEMMA v0.97^51^. We focussed this analysis on lead *cis*-pQTLs to identify interactions of genetic variants with BMI SDS. NPX protein levels were rank-inverse normal transformed and linearly adjusted for the year of blood sample collection, age, sex and Olink batch, and genotype batch was added as a covariate in the analysis.

### Comparison with the UK Biobank

We compared pQTLs in 3,853 individuals from the HOLBAEK Study with pQTLs in 3,853 individuals from the UK Biobank. Association analyses were performed using REGENIE v3^60^ on genotype dosages with similar covariates used in both cohorts. We included study-wide significant lead *cis*-pQTLs in the HOLBAEK Study in the comparison (n=90) and only those with a matching SNP in the UK Biobank were retained (n=75; 15 removed). To estimate the effect of sample size, GWAS software, and covariates on this analysis, we also included pQTL summary statistics for 34,557 individuals from the UK Biobank (synapse.org, under Project SynID: syn51364943) and summary statistics generated with GEMMA from the meta-analyses of obesity clinic and population cohorts of the HOLBAEK Study (n=3,853) in the comparison. We lifted variants from Hg19 to Hg38 with the liftover R package^61^ and used the bigsnpr R package for harmonization^62^.

We ran downsampled and obesity-dependent pQTL analyses using the UK Biobank Research Analysis Platform on DNAnexus. We included participants with European ancestry with measurements at baseline and excluded consortium-selected participants and participants with a positive COVID-19 status (Data-Fields 22006, 30903 and Resource 1016). For the downsampling analysis, we included individuals who had measurements for most of the plasma proteins (>2,900 proteins; n=19,969) and we randomly sampled 3,853 individuals (55.85% females) to match the sample size and male-to-female ratio of the HOLBAEK Study. For the obesity-dependent analysis, we separated participants from the UK Biobank into two groups: 1) participants with a BMI ≥30 (n=8,803; 51.90% female; median age 59 years, Q1– Q3: 52–64), as a proxy for obesity, and 2) a control group with a BMI <30 (n=8,803; 51,90% female; median age 58, Q1–Q3: 50–63).

For the age-dependent comparison, pQTL analyses were run with REGENIE v3^60^ using called genotypes in step 1 and imputed genotype dosages in step 2. In the UK Biobank, we rank-inverse normalized protein levels and included age, age^2^, sex, age × sex, age^2^ × sex, Olink batch, UK Biobank assessment centre, genotype batch, and the first 20 genetic principal components (Data-Fields 21022, 31, 30901, 54, 22000, and 22009) as covariates for consistency with the pQTL analysis previously performed by Sun et al.^1^. In the HOLBAEK Study, we rank-inverse normalized protein levels, adjusted them for the Olink panel batch, and included age, age^2^, sex, age × sex, age^2^ × sex, year of blood sample collection, cohort status, genotyping batch, and 10 genetic principal components as covariates in the association analysis. We ran REGENIE with block sizes of 1000 and 400 in step 1 and 2. Genotype quality control was performed with PLINK2^63^ and included removing individuals with more than 5% missing genotypes and genetic variants with a genotyping rate below 95%. We included autosomal variants with minor allele frequency > 1%, Hardy-Weinberg equilibrium test P-value > 10^-6^ and imputation score > 0.7. Heterogeneity between pQTLs of the HOLBAEK Study and the UK Biobank GWAS was assessed with METAL^52^.

### Olink platform comparison

We compared the plasma proteomic measurements between the Olink Target 96 and Olink Explore 3072 platforms in 739 individuals without signs of CAD, determined by coronary computed tomography angiography (CCTA) from the Dan-NICAD cohort^27,28^. For the comparison, 178 proteins from the Olink Target 96 Inflammation and CVD-II panels were preferably matched to proteins in the Explore 3072 Inflammation and Cardiometabolic subpanels. Protein NPX values were rank-inverse normalized, adjusted for sample collection box ID to reflect time-of-year of sample collection, followed by another rank-inverse normalization. For each protein, the Pearson correlation between the Olink Target and Olink Explore platforms was reported (Table S8).

### Comparison of protein levels in obesity

We compared plasma levels of proteins with obesity-dependent *cis*-pQTLs in the obesity clinic- and population-based cohorts in the HOLBAEK Study (Table S6). We further compared levels of the same proteins in individuals with a BMI ≥ 30 kg/m^2^ and a control group with BMI < 30 kg/m^2^ in the UK Biobank. We rank-inverse normal transformed protein levels and adjusted the proteins for the same set of covariates used in the genetic association analysis, excluding genetic covariates such as genotype batch and genetic principal components.

### One-sample MR analysis in the HOLBAEK Study

One-sample MR analysis was performed using individual-level data from the HOLBAEK Study, applying the two-stage least squares approach implemented in the ivreg R package. We only included individuals from the population-based cohort, due to the risk of selection bias in the obesity clinic cohort. We further excluded related individuals (1,825 retained). We limited the one-sample MR analysis to 90 proteins with study-wide significant *cis*-pQTLs identified in the meta-analysis of the HOLBAEK Study and 21 cardiometabolic traits including traditional lipids, glycemic traits, inflammatory markers, liver-related markers, metabolic hormones, and blood pressure. The study-wide *cis*-pQTL explaining the largest proportion of variance in protein levels was used as the instrumental variable. In stage 1, protein levels were regressed on the instrumental variable, with age, sex, BMI SDS, year of blood sample collection, genotype batch, and the first ten genetic principal components included as covariates. In stage 2, the predicted protein levels were regressed on cardiometabolic traits, with the same covariates as in stage 1. Protein levels were adjusted for the Olink batch and rank-inverse normalized before the analysis. Cardiometabolic traits were log-transformed. Transformed protein levels and cardiometabolic traits were then centered around 0 and scaled to a standard deviation of 1. We tested for weak instruments with the sensitivity analysis included in the ivreg R package. Collection of cardiometabolic traits in the HOLBAEK Study was previously described elsewhere^12^.

## Acknowledgments

We thank all volunteers and their parents who participated in the HOLBAEK Study. The HOLBAEK study is registered at ClinicalTrials.gov (NCT00928473). This research has been conducted using the UK Biobank Resource under Application Number 32683 and uses data provided by patients and collected by the NHS as part of their care and support. Data on childhood body mass index have been contributed by the EGG Consortium and have been downloaded from www.egg-consortium.org.

## Author contributions

Roman Thielemann had full access to all of the data in the study and took full responsibility for the integrity of the data and the accuracy of the data analysis.

Concept and Design: RT, SES, YH, JCH, SR, TH

Acquisition, analysis, or interpretation of data: RT, SES, YH, LAH, JFG, PLM, PDR, AIP, LVA, CEF, MT, AK, SR, JCH, TH

Drafting of the manuscript: RT, SES, YH, TH

Critical revision of the manuscript for important intellectual content: RT, SES, YH, LAH, JFG, PLM, PDR, AIP, LVA, CEF, PLM, PDR, MT, AK, SR, JCH, TH

Statistical analysis: RT, SES, YH Obtained funding: JCH, TH, MT, AK Supervision: SR, TH

## Data availability

Results from statistical analysis are available in the Supplementary Tables 1–12. GWAS summary statistics will be made available upon publication.

## References

1. Sun, B. B. et al. Plasma proteomic associations with genetics and health in the UK Biobank. Nature 622, 329–338 (2023).

2. Folkersen, L. et al. Genomic and drug target evaluation of 90 cardiovascular proteins in 30,931 individuals. Nat Metab 2, 1135–1148 (2020).

3. Zhao, J. H. et al. Genetics of circulating inflammatory proteins identifies drivers of immune-mediated disease risk and therapeutic targets. Nat. Immunol. 24, 1540–1551 (2023).

4. Eldjarn, G. H. et al. Large-scale plasma proteomics comparisons through genetics and disease associations. Nature 622, 348–358 (2023).

5. Sun, B. B. et al. Genomic atlas of the human plasma proteome. Nature 558, 73–79 (2018).

6. Pietzner, M. et al. Mapping the proteo-genomic convergence of human diseases. Science 374, eabj1541 (2021).

7. Karhunen, V. et al. The interplay between inflammatory cytokines and cardiometabolic disease: bi-directional mendelian randomisation study. BMJ Med 2, e000157 (2023).

8. Thareja, G. et al. Differences and commonalities in the genetic architecture of protein quantitative trait loci in European and Arab populations. Hum. Mol. Genet. 32, 907–916 (2023).

9. Xu, F. et al. Genome-wide genotype-serum proteome mapping provides insights into the cross-ancestry differences in cardiometabolic disease susceptibility. Nat. Commun. 14, 896 (2023).

10. Said, S., et al. Ancestry diversity in the genetic determinants of the human plasma proteome and associated new drug targets. *bioRxiv* (2023) doi:10.1101/2023.11.13.23298365.

11. Niu, L. et al. Plasma proteome variation and its genetic determinants in children and adolescents. Nat. Genet. 1–12 (2025) doi:10.1038/s41588-025-02089-2.

12. Stinson, S. et al. Identification of modifiable plasma protein markers of cardiometabolic risk in children and adolescents with obesity. medRxiv 2025.02.25.25322850 (2025) doi:10.1101/2025.02.25.25322850.

13. Koprulu, M. et al. Similar and different: systematic investigation of proteogenomic variation between sexes and its relevance for human diseases. Genetic and Genomic Medicine (2024).

14. Wingo, A. P. et al. Sex differences in brain protein expression and disease. Nat. Med. 29, 2224–2232 (2023).

15. Hillary, R. F. et al. Systematic discovery of gene–environment interactions underlying the human plasma proteome in UK Biobank. bioRxiv (2023) doi:10.1101/2023.10.26.23297604.

16. Helgeland, Ø. et al. Characterization of the genetic architecture of infant and early childhood body mass index. Nat Metab 4, 344–358 (2022).

17. Smith, J. G. & Gerszten, R. E. Emerging affinity-based proteomic technologies for large-scale plasma profiling in cardiovascular disease. Circulation 135, 1651–1664 (2017).

18. Soták, M., Clark, M., Suur, B. E. & Börgeson, E. Inflammation and resolution in obesity. Nat. Rev. Endocrinol. 21, 45–61 (2025).

19. Singer, K. & Lumeng, C. N. The initiation of metabolic inflammation in childhood obesity. J. Clin. Invest. 127, 65–73 (2017).

20. Berenson, G. S. et al. Association between multiple cardiovascular risk factors and atherosclerosis in children and young adults. The Bogalusa Heart Study. N. Engl. J. Med. 338, 1650–1656 (1998).

21. Raitakari, O., Pahkala, K. & Magnussen, C. G. Prevention of atherosclerosis from childhood. Nat. Rev. Cardiol. 19, 543–554 (2022).

22. Pang, Y. et al. Associations of adiposity, circulating protein biomarkers, and risk of major vascular diseases. JAMA Cardiol. 6, 276–286 (2021).

23. Lausten-Thomsen, U. et al. Reference values for serum total adiponectin in healthy non-obese children and adolescents. Clin. Chim. Acta 450, 11–14 (2015).

24. Holm, J.-C. et al. Chronic care treatment of obese children and adolescents. Int. J. Pediatr. Obes. 6, 188–196 (2011).

25. Vogelezang, S. et al. Novel loci for childhood body mass index and shared heritability with adult cardiometabolic traits. PLoS Genet. 16, e1008718 (2020).

26. Feizi, A. & Ray, K. otargen: GraphQL-based R package for tidy data accessing and processing from Open Targets Genetics. Bioinformatics 39, (2023).

27. Nissen, L. et al. Danish study of Non-Invasive testing in Coronary Artery Disease (Dan-NICAD): study protocol for a randomised controlled trial. Trials 17, 262 (2016).

28. Nissen, L. et al. Diagnosing coronary artery disease after a positive coronary computed tomography angiography: the Dan-NICAD open label, parallel, head to head, randomized controlled diagnostic accuracy trial of cardiovascular magnetic resonance and myocardial perfusion scintigraphy. Eur. Heart J. Cardiovasc. Imaging 19, 369–377 (2018).

29. Goulding, N. et al. Inflammation proteomics datasets in the ALSPAC cohort [version 2; peer review: 2 approved]. Wellcome Open Research 7, (2024).

30. Arend, W. P., Malyak, M., Guthridge, C. J. & Gabay, C. Interleukin-1 receptor antagonist: role in biology. Annu. Rev. Immunol. 16, 27–55 (1998).

31. Ghanbari, M. et al. Interleukin-1 in obesity-related low-grade inflammation: From molecular mechanisms to therapeutic strategies. Int. Immunopharmacol. 96, 107765 (2021).

32. Walsh, M. C. & Choi, Y. Biology of the TRANCE axis. Cytokine Growth Factor Rev. 14, 251–263 (2003).

33. Lacey, D. L. et al. Osteoprotegerin ligand is a cytokine that regulates osteoclast differentiation and activation. Cell 93, 165–176 (1998).

34. Wong, B. R. et al. TRANCE Is a Novel Ligand of the Tumor Necrosis Factor Receptor Family That Activates c-Jun N-terminal Kinase in T Cells*. J. Biol. Chem. 272, 25190– 25194 (1997).

35. Body, J.-J. et al. A phase I study of AMGN-0007, a recombinant osteoprotegerin construct, in patients with multiple myeloma or breast carcinoma related bone metastases. Cancer 97, 887–892 (2003).

36. Bekker, P. J. et al. The effect of a single dose of osteoprotegerin in postmenopausal women. J. Bone Miner. Res. 16, 348–360 (2001).

37. Spohn, G. et al. Protection against osteoporosis by active immunization with TRANCE/RANKL displayed on virus-like particles. J. Immunol. 175, 6211–6218 (2005).

38. Matsumoto, T. & Endo, I. RANKL as a target for the treatment of osteoporosis. J. Bone Miner. Metab. 39, 91–105 (2021).

39. Irvine, K. M. et al. Serum matrix metalloproteinase 7 (MMP7) is a biomarker of fibrosis in patients with non-alcoholic fatty liver disease. Sci. Rep. 11, 2858 (2021).

40. Huang, C.-C. et al. Matrilysin (MMP-7) is a major matrix metalloproteinase upregulated in biliary atresia-associated liver fibrosis. Mod. Pathol. 18, 941–950 (2005).

41. Bauer, Y. et al. MMP-7 is a predictive biomarker of disease progression in patients with idiopathic pulmonary fibrosis. ERJ Open Res. 3, 00074–02016 (2017).

42. Cai, B. et al. Macrophage MerTK promotes liver fibrosis in nonalcoholic steatohepatitis. Cell Metab. 31, 406–421.e7 (2020).

43. Iimuro, Y. et al. Delivery of matrix metalloproteinase-1 attenuates established liver fibrosis in the rat. Gastroenterology 124, 445–458 (2003).

44. Ligthart, S. et al. En-rage: A novel inflammatory marker for incident coronary heart disease. Arterioscler. Thromb. Vasc. Biol. 34, 2695–2699 (2014).

45. Oesterle, A. & Bowman, M. A. H. S100A12 and the S100/calgranulins: Emerging biomarkers for atherosclerosis and possibly therapeutic targets: Emerging biomarkers for atherosclerosis and possibly therapeutic targets. Arterioscler. Thromb. Vasc. Biol. 35, 2496–2507 (2015).

46. Suhre, K. et al. Nanoparticle enrichment mass-spectrometry proteomics identifies protein-altering variants for precise pQTL mapping. Nat. Commun. 15, 989 (2024).

47. Virolainen, S. J., VonHandorf, A., Viel, K. C. M. F., Weirauch, M. T. & Kottyan, L. C. Gene-environment interactions and their impact on human health. Genes Immun. 24, 1– 11 (2023).

48. Zhong, W. et al. Next generation plasma proteome profiling to monitor health and disease. Nat. Commun. 12, 2493 (2021).

49. Stinson, S. E. et al. High Plasma Levels of Soluble Lectin-like Oxidized Low-Density Lipoprotein Receptor-1 Are Associated With Inflammation and Cardiometabolic Risk Profiles in Pediatric Overweight and Obesity. J. Am. Heart Assoc. 12, e8145 (2023).

50. McCarthy, S. et al. A reference panel of 64,976 haplotypes for genotype imputation. Nat. Genet. 48, 1279–1283 (2016).

51. Zhou, X. & Stephens, M. Genome-wide efficient mixed-model analysis for association studies. Nat. Genet. 44, 821–824 (2012).

52. Willer, C. J., Li, Y. & Abecasis, G. R. METAL: fast and efficient meta-analysis of genomewide association scans. Bioinformatics 26, 2190–2191 (2010).

53. Purcell, S. et al. PLINK: a tool set for whole-genome association and population-based linkage analyses. Am. J. Hum. Genet. 81, 559–575 (2007).

54. Marshall, W. A. & Tanner, J. M. Variations in the pattern of pubertal changes in boys. Arch. Dis. Child. 45, 13–23 (1970).

55. Marshall, W. A. & Tanner, J. M. Variations in pattern of pubertal changes in girls. Arch. Dis. Child. 44, 291–303 (1969).

56. Juul, A. et al. Pubertal development in Danish children: comparison of recent European and US data. Int. J. Androl. 29, 247–55; discussion 286–90 (2006).

57. Viechtbauer, W. Conducting meta-analyses in R with the metafor package. Journal of Statistical Software vol. 36 1–48 Preprint at 10.18637/jss.v036.i03 (2010).

58. McLaren, W. et al. The Ensembl Variant Effect Predictor. Genome Biol. 17, 122 (2016).

59. Durinck, S., Spellman, P. T., Birney, E. & Huber, W. Mapping identifiers for the integration of genomic datasets with the R/Bioconductor package biomaRt. Nat. Protoc. 4, 1184–1191 (2009).

60. Mbatchou, J. et al. Computationally efficient whole-genome regression for quantitative and binary traits. Nat. Genet. 53, 1097–1103 (2021).

61. Bioconductor Package Maintainer. liftOver: Changing genomic coordinate systems with rtracklayer::liftOver. Preprint at https://www.bioconductor.org/help/workflows/liftOver/ (2023).

62. Privé, F., Aschard, H., Ziyatdinov, A. & Blum, M. G. B. Efficient analysis of large-scale genome-wide data with two R packages: bigstatsr and bigsnpr. Bioinformatics 34, 2781–2787 (2018).

63. Chang, C. C. et al. Second-generation PLINK: rising to the challenge of larger and richer datasets. Gigascience 4, 7 (2015).

