## Supplemental Figures 1-7 for "Obesity- and age-dependent genetic regulation of the plasma proteome in children and adolescents"

### **Supplementary figures**

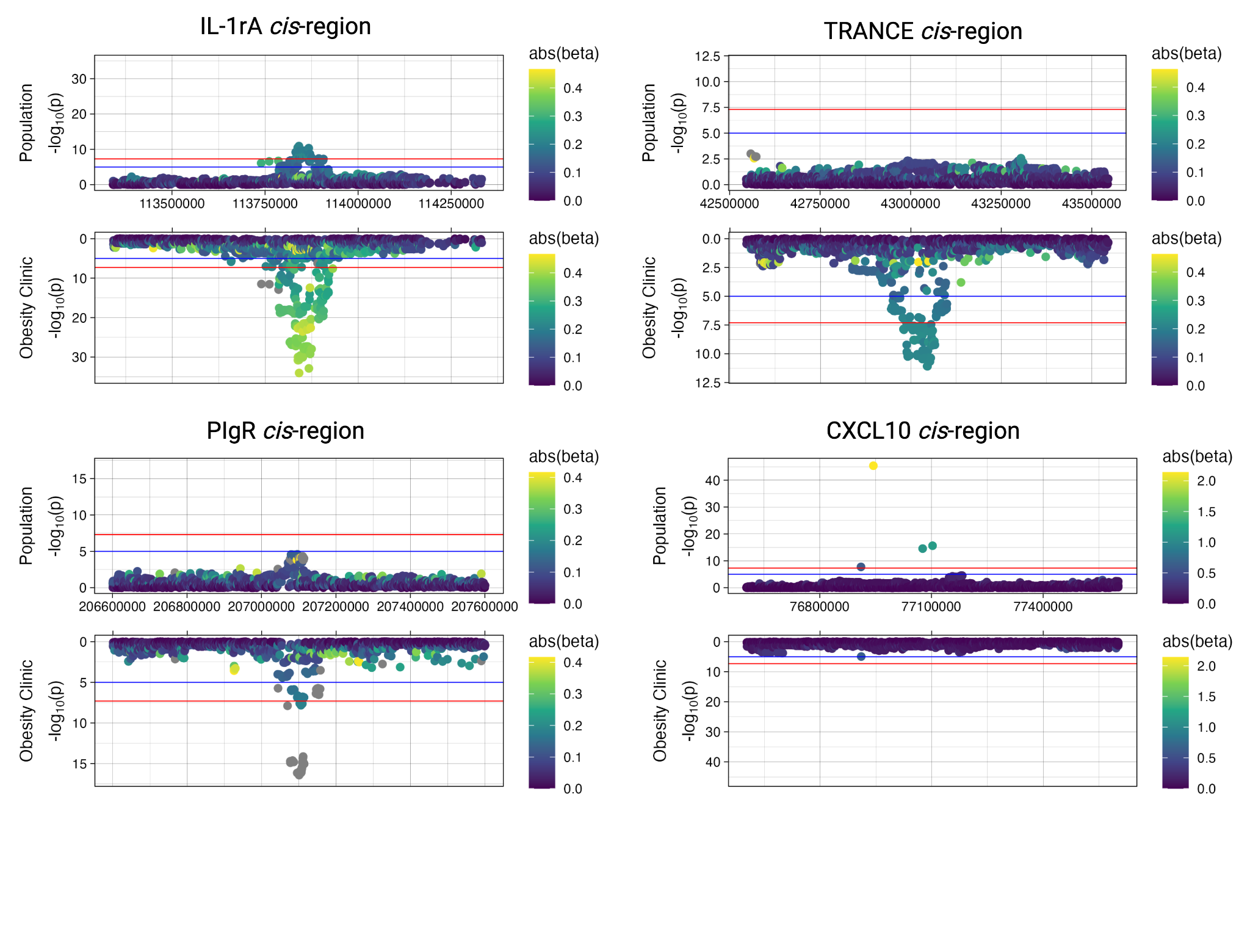

**Supplemental Figure 1: *Cis*-region zoom plot of obesity-dependent pQTLs.** GWAS signals are shown for *cis*-loci in the population cohort (upper Manhattan plot) and obesity clinic cohort (lower Manhattan plot). Suggestive significance (10^-5^; blue) and genome-wide significance (5✕10^-8^; red) thresholds are highlighted. Variants are colored by their absolute beta values.

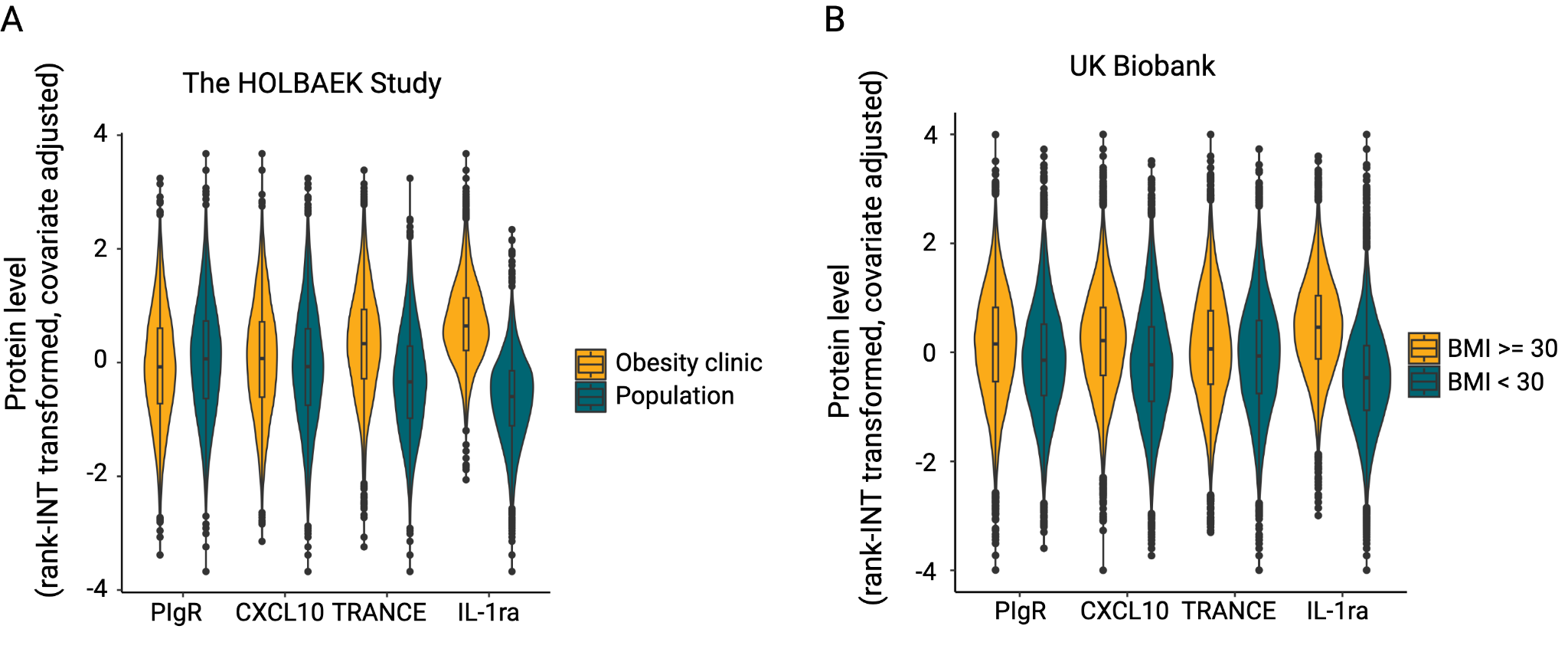

**Supplemental Figure 2: Plasma levels of proteins with obesity-dependent *cis*-pQTLs.** PIgR, CXCL10, TRANCE, and IL1ra levels are shown in A) The HOLBAEK Study and B) the UK Biobank. Protein levels were rank-inverse normal transformed and adjusted for covariates.

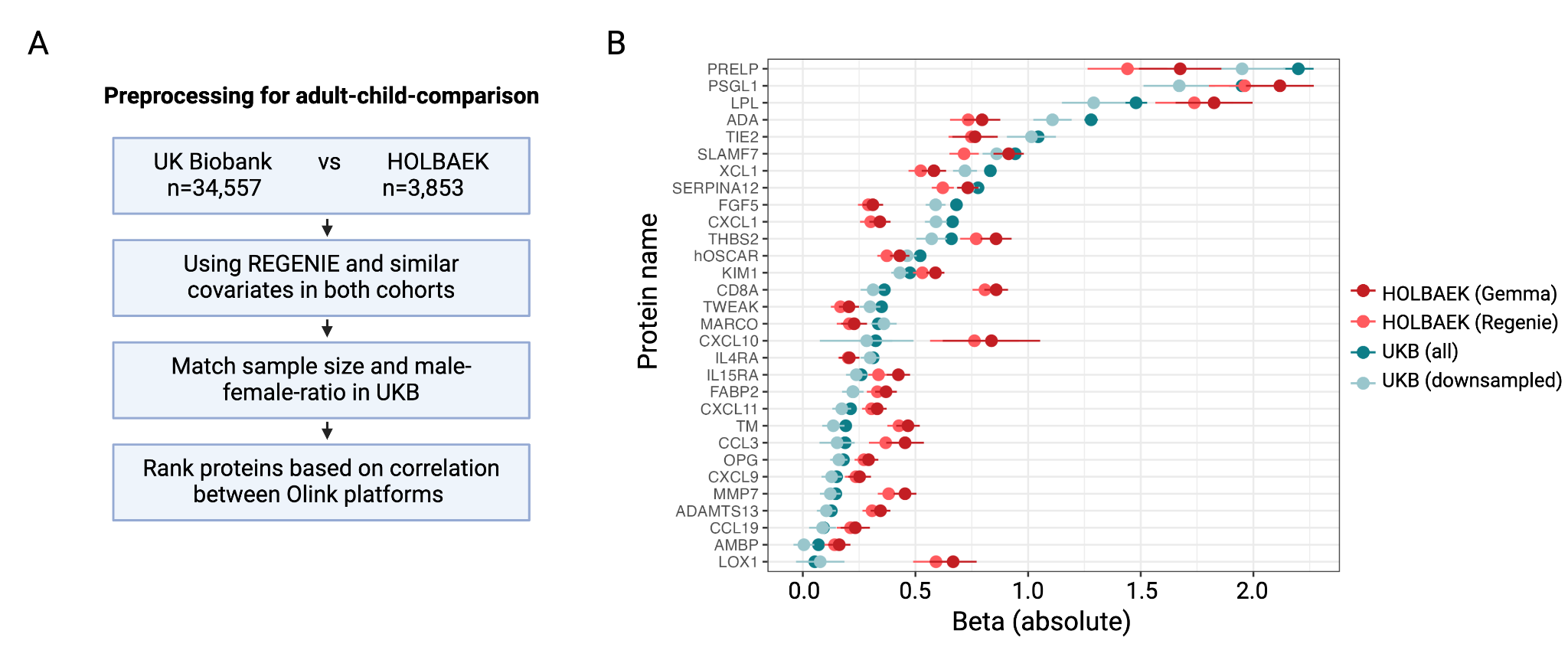

**Supplemental Figure 3: Effects of downsampling and rerunning GWAS with REGENIE on *cis*-pQTLs.** A) Preprocessing steps to address potential sources of bias such as sample size, covariates used in the association analysis, and GWAS methods, and differences in proteomic measurements. B) Beta values differed systematically when using different GWAS approaches in the HOLBAEK Study (GEMMA or REGENIE) and for different sample sizes in the UK Biobank (n=34,557 or n=3,853). Top 30 proteins with age-dependent differences are shown.

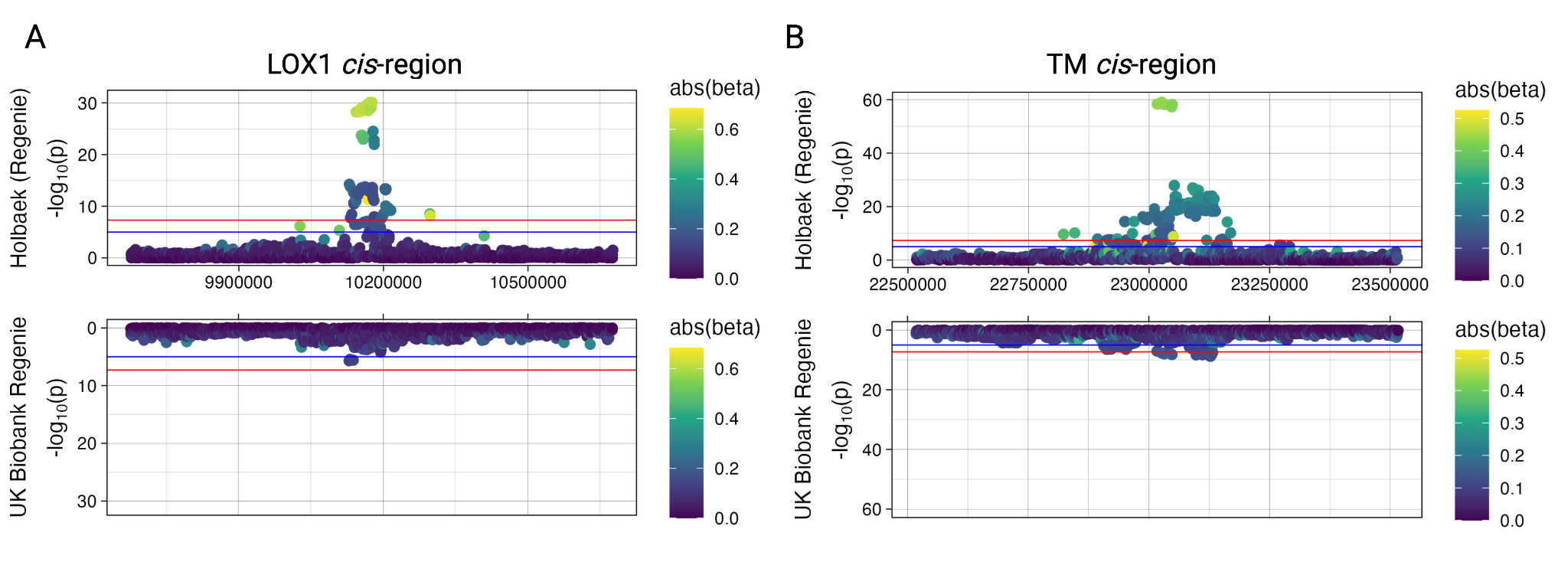

**Supplemental Figure 4: Zoom plot for cis-pQTLs of LOX1 and TM.** Zoom plots of *cis*-regions for A) LOX1 and B) TM reveals age-dependent differences between the HOLBAEK Study and the UK Biobank. Genomic coordinates in this figure refer to genome-build GRCh38. Suggestive significance (10^-5^, blue) and genome-wide significance (5 × 10^-8^, red) thresholds are highlighted.

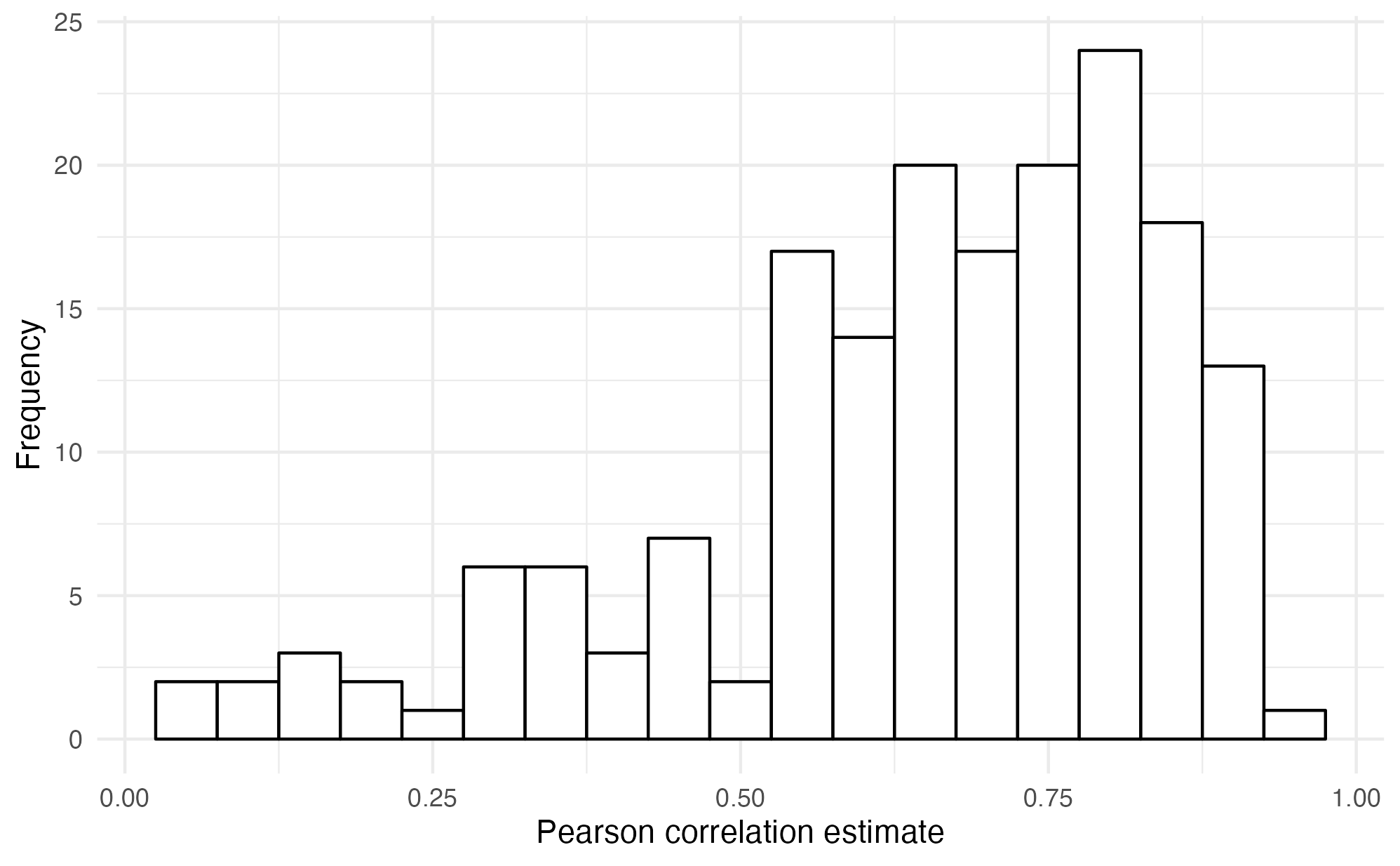

**Supplemental Figure 5: Correlation of Olink protein measurements across the Target 96 and Explore 3072 platforms.** Histogram of Pearson correlation estimates for 178 proteins in 739 individuals of the Dan-NICAD cohort. The median Pearson correlation estimate was 0.69 (Q1–Q3: 0.57–0.80).

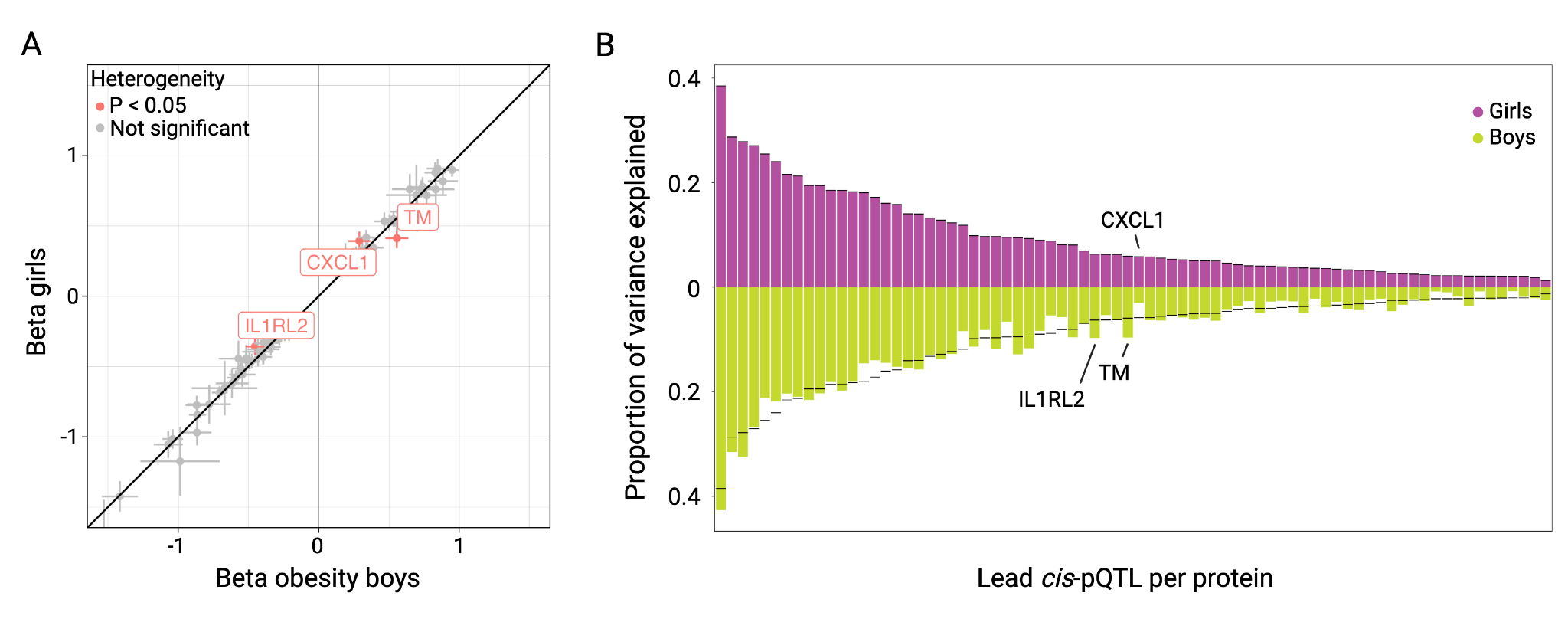

**Supplemental Figure 6: *Cis*-pQTLs differences in boys and girls.** A) Differences in beta values of plasma *cis*-pQTLs between boys and girls. We found nominally significant evidence for sex-dependent beta value differences in pQTLs of CXCL1, IL1RL2, and TM. B) *Cis*-pQTLs explained a similar proportion of variance in protein levels across cohorts. Proteins with nominal evidence for heterogeneity were annotated.

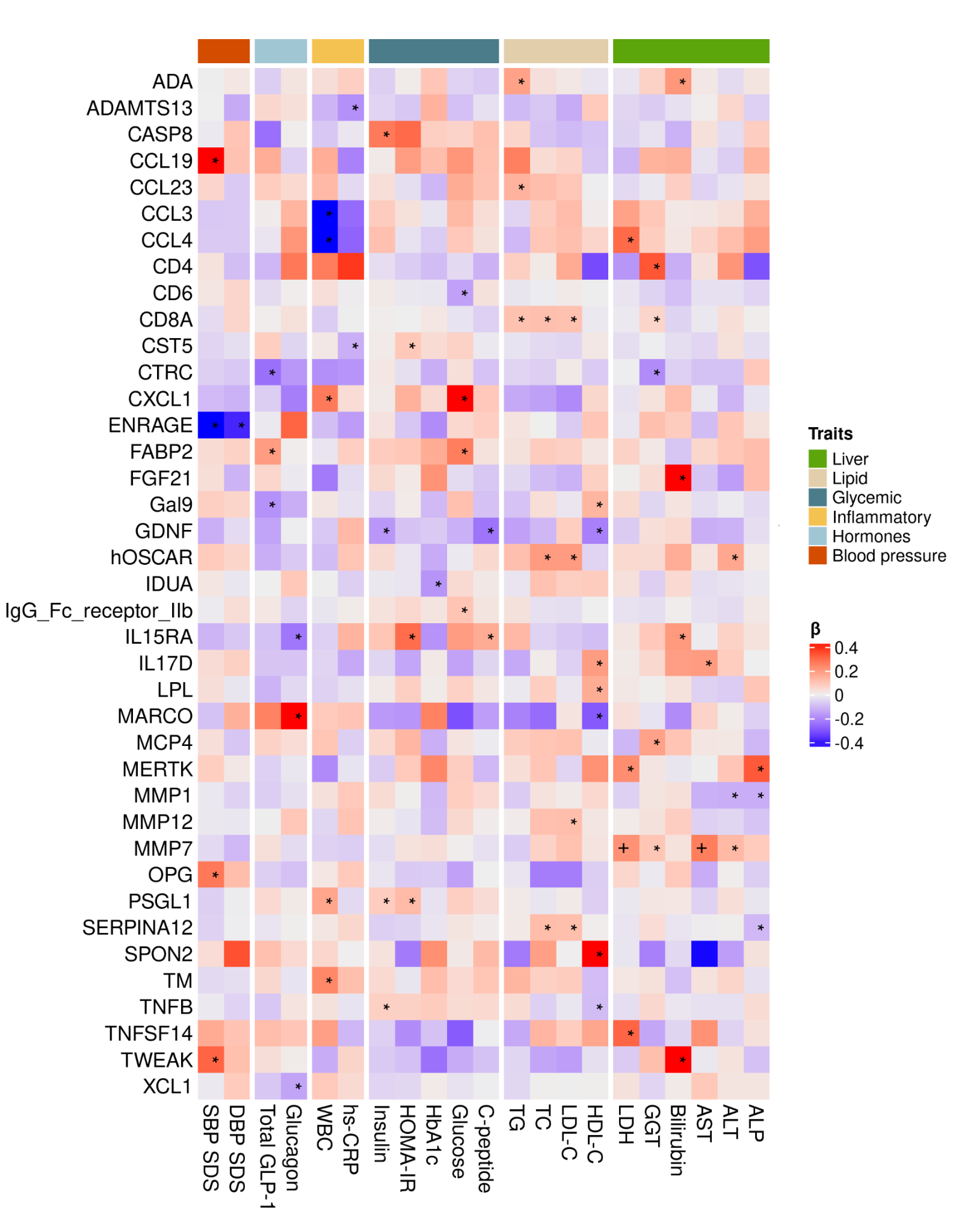

**Supplemental Figure 7: Heatmap of one-sample MR results for proteins and cardiometabolic traits.** Only proteins with nominally significant association are shown. Significance levels are labeled with * for nominally significance (P-value < 0.05) and + for FDR significance (FDR < 5%).
